# Antipsychotic Drugs and the Risk of Breast Cancer

**DOI:** 10.1101/2021.06.06.21258408

**Authors:** Tahir Rahman, John M. Sahrmann, Margaret A. Olsen, Katelin B. Nickel, J. Phillip Miller, Cynthia Ma, Richard A. Grucza

## Abstract

**Objective:** Antipsychotic drugs are well established to alter circulating prolactin levels by blocking dopamine D-2 receptors in the pituitary. Prolactin activates many genes important in the development of breast cancer. The aim of this study was to evaluate the risk of breast cancer in women exposed to antipsychotic drugs, stratified by prolactin elevating potential (high, mid, and low), compared to women taking anticonvulsants and/or lithium.

**Methods:** The IBM MarketScan Commercial and Medicaid Databases were used to establish a large, observational cohort of women taking antipsychotics drugs compared to control drugs. Invasive breast cancer was identified using diagnostic codes. Bivariable and multivariable Cox proportional hazards models were used to evaluate the risk of breast cancer by antipsychotic drug exposure, both as pooled antipsychotics and by prolactin specific categories.

**Results:** A total of 2,708 (0.2%) cases of invasive breast cancer were identified among 1,562,839 women. Exposure to antipsychotics with high prolactin elevating potential was associated with a 23% increased risk of breast cancer (aHR 1.23; 95% CI, 1.11-1.35), whereas mid and low prolactin categories of antipsychotics were not significant.

**Conclusion:** In the largest study of antipsychotics taken by women, a modest risk between antipsychotic drug use and the risk for breast cancer was observed, with a differential higher association with high prolactin elevating drugs. Residual confounding factors included incomplete information on parity, race and socioeconomic status, and differential outpatient visits. Clinicians should consider monitoring serum prolactin levels and adopting vigilant mammography screening practices, especially in older women taking category one antipsychotics.

## Introduction

The incidence of breast cancer in the United States is in excess of 250,000 cases per year. Approximately 12.3% of women will be diagnosed with breast cancer at some point during their lifetime (1). Endocrine evidence suggests that prolactin (PRL), in addition to other factors, plays an important role in the development of breast tumorigenesis. Prolactin receptor (PRLr) is overexpressed by 95% of breast cancers and activates many genes responsible for proliferation and metastatic spread of breast cancer through PRL-PRLr-JAK2-STAT5 signaling (2, 3). Many antipsychotic drugs are well known to produce elevated serum prolactin and can induce breast neoplasms in rodent studies, as mentioned in their package inserts (2-4). Hyperprolactinemia from antipsychotic drug use is caused by blockade of dopamine (D-2) receptors in the tuberoinfundibular tract by removing inhibitory influence on lactotroph cells in the anterior pituitary (2, 15). Common side effects in humans include amenorrhea, oligomenorrhea, osteoporosis, gynecomastia, early telarche, sexual dysfunction, and infertility (16). High breast tissue density is an intermediate phenotype for breast cancer and is also positively associated with prolactin levels. Mouse models that closely mimic human breast cancer initiation, have shown that hyperprolactinemia-inducing antipsychotics accelerate early lesion progression to cancerous cells, thus raising concerns for older women that are more likely to have precancerous lesions (3).

Human studies in the general population have addressed the associations between prolactin levels and risk of developing breast cancer which are considered modest (risk ratios ranging from 0.70 to 1.9 for premenopausal women and from 0.76 to 2.03 for postmenopausal women) (2, 5, 6). The Nurses’ Health Study, a robust prospective cohort study, showed that both pre- and postmenopausal women who are in the top quartile of serum prolactin levels have higher risk of developing breast cancer compared to those in the lowest quartile. Antipsychotic drugs have also been separately studied for an association with breast cancer and found to have mixed associations (4-6, 10-15). An often cited observational study by Wang, et al examined over 100,000 women and compared 52,819 women on dopamine antagonists to 55,289 women not treated with them. Women who used antipsychotic dopamine antagonists had a 16% greater risk (adjusted HR 1.16, 95% CI, 1.07–1.26) of developing breast cancer compared to never users, with a dose–response relationship between larger cumulative dosages and greater risk (4). Another population based cohort study in Taiwan stratified drugs by prolactin elevating potential and found higher rates of breast cancer in women with schizophrenia treated with risperidone, paliperiodone or misulpride. (adjusted HR 1.96, 95% CI, 1.36-2.82) (11). However, systematic reviews of the literature as well as expert panels have concluded that antipsychotics pose little or no risk of breast cancer (6, 16), while others have advised caution when prescribing prolactin elevating drugs to women with a previously detected breast cancer (2, 17).

One problem that the current classification of antipsychotics has is the potential to cause confusion because the terms “first and second generation” were arbitrarily assigned to many antipsychotics drugs depending on their first release to the market and potential to cause extrapyramidal side effects, and not based solely on propensity to raise prolactin (20, 21). Normal prolactin levels are around 20-25 ng/ml. Typical or “first generation” antipsychotics, as well as risperidone, and paliperidone have the highest propensity to raise prolactin (45-to >100 ng/ml), whereas “atypical” antipsychotics are associated with more modest elevations, while aripirazole/ brexpriprazole are not associated with it (2, 19, 22-29). A more concise way would be to categorize drugs according to known prolactin elevating properties (2, 19). Utilizing such stratification, we conducted the largest epidemiologic study to date investigating the risk of breast cancer and antipsychotics from an administrative claims database of over 150 million Americans. We hypothesized that women prescribed prolactin elevating antipsychotic drugs would have higher risk of breast cancer compared to women taking anticonvulsants or lithium as the comparator group. We further hypothesized that relative to control drugs, the risk of breast cancer would follow the order of high, mid and low prolactin elevating categories of antipsychotics.

## Methods

### Data Sources

We performed a large, retrospective, observational cohort study of breast cancer risk in antipsychotic exposed female patients 18–64 years of age using administrative claims data from the IBM® MarketScan® Commercial and Multi-State Medicaid Databases.^1^ Persons 65 years of age and older are not included in this database as they would likely be covered by Medicare and therefore not have complete data. The MarketScan Commercial Database includes medical/ outpatient prescription drug claims from 150 million privately insured persons, and the Medicaid database includes similar information from 20 million Medicaid insured persons. Variables were defined using information (e.g., diagnosis codes, procedure codes) on medical/ outpatient prescription drug claims submitted for reimbursement. The databases contain no patient identifiers and the study was considered exempt by the Washington University in St. Louis School of Medicine, Human Research Protection Office.

### Study population

We identified women between the ages of 18 to 64 with a outpatient prescription drug claim with days’ supply > 0 days for an antipsychotic, anticonvulsant, or lithium from 1/1/2006 through 6/30/2016 (1/1/2011 through 6/30/2016 from the Medicaid Multi-State database). Supplemental Table 1 contains the generic drug names for each category of drug in the study. We required at least six months of continuous medical and prescription drug insurance enrollment to identify baseline covariates. Each woman was assigned an index date equal to six months after the start of insurance enrollment for those with a claim for a study drug during that time or the date of the first claim for a study drug for women without a prescription for a study drug during the first six months of enrollment.

### Quantification of Exposure to Antipsychotics and Control Drugs

We separated antipsychotic drugs into three categories (category 1 highest to category 3 lowest) based on their propensity to elevate prolactin. (Supplemental Table 1). These categories were derived *a priori* from previously published antipsychotic clinical guides (2, 19). We chose anticonvulsants and lithium as comparator drugs since they are not known to raise prolactin and are prescribed to patients with a serious mental illness, and thus could mitigate potential confounders (30-33). Exposure within the drug categories was measured using drug-specific defined daily dose (DDD), a quantity developed by the World Health Organization’s Collaborating Centre for Drug Statistics Methodology (34). The strength, quantity, and days’ supply listed on each claim were used to compute per-day DDD, with upper limits applied based on typical prescribing patterns determined by one author (TR, Supplemental Table 1). To determine each woman’s cumulative exposure to each category of drug, we summed DDDs across all claims beginning with the first fill of a study drug and ending with the first outcome or censoring event (i.e., end of enrollment, six years after index, or evidence of prevalent disease [see below]). We then divided cumulative exposure by the length of this time period to obtain the average exposure during the observed treatment time. The average DDD per day of observed treatment was calculated as the cumulative DDD/total days of observed treatment period. For example, a woman with prescriptions for 300 mg/day of chlorpromazine (category 1 medication) covering every day of her study observation would have an average DDD of 1.00 for chlorpromazine (300 mg defined by the WHO as the average daily maintenance dose). If the same woman had prescriptions for the same dose of chlorpromazine for 10% of her study observation time, the average DDD for chlorpromazine would be 0.10 per day. Women treated solely with prochlorperazine were not included, since it is also an antiemetic, but it was included in the calculation of the total DDD exposure to Category 1 antipsychotics with other study drug exposures.

### Identification of Invasive Breast Cancer

We identified invasive breast cancer using hierarchical definitions based on the quality of available evidence from diagnosis/ procedure codes (Supplemental Table 2). We first excluded likely prevalent cases indicated by a prescription drug claim for tamoxifen or a diagnosis of breast cancer or history of breast cancer on any claim prior to the index date. Invasive breast cancer was identified primarily by International Classification of Diseases, 9th/10th edition, Clinical Modification, (ICD-9/10) codes for invasive breast cancer on a claim with a Current Procedural Terminology (CPT-4) code for surgical pathology microscopic examination (Supplemental Table 2), indicating pathologic verification (35). If this was not present, a diagnosis of invasive breast cancer on an inpatient facility claim or on at least two provider/outpatient claims separated by 30-180 days was required, with the later of the two dates for outpatient diagnoses used as the date of invasive cancer. For potential cases of invasive breast cancer without pathologist confirmation, evidence of surgical treatment (mastectomy or breast-conserving surgery within one month before through six months after diagnosis), or chemotherapy (within six months after diagnosis, chemotherapy administration coded for invasive breast cancer) was required to establish the diagnosis of invasive breast cancer. Diagnoses without associated surgical or chemotherapeutic treatment were assumed to represent prevalent/ unverified cases and were treated as censoring events.

### Identification of Covariates

Covariates of interest were identified using ICD-9/10 diagnosis codes and ICD-9/10 procedure and CPT-4 codes from inpatient and outpatient claims, and prescription drug claims for some covariates. Baseline covariates assessed using all available claims (36) prior to the index date included potential risk factors for invasive breast cancer, including benign breast disease, smoking and smoking-related diseases, diabetes, alcohol abuse, obesity, and additional proxies for obesity, including sleep apnea, gastroesophageal reflux disease, lipodystrophy, and other lipidemias (Supplemental Table 2). The additional proxies for obesity were included in the definition of obesity due to the low sensitivity of coding for obesity in claims data, including in breast cancer patients (37). We used prescription drug claims to identify lipid-lowering medications, statins, smoking cessation therapy, and antidiabetic agents, to improve sensitivity of their identification. Hormone replacement therapy was identified using prescription drug claims in the therapeutic class ‘Estrogens & Combinations’, categorized as estrogen only vs estrogen plus progesterone. Hormone replacement therapy was restricted to women aged 50 years and older or younger women with a diagnosis of acquired absence of genital organs. Preexisting benign breast disease was identified by diagnosis or procedure codes (Supplemental Table 2), but was not counted if it was first coded within 180 days of the date of invasive breast cancer, since it was most likely concurrent with the invasive breast cancer. For all variables defined by diagnosis or procedure codes, the absence of codes for the condition in the claims data was interpreted as lack of the condition of interest (and the variable thus coded 0), as is typically done in analyses of claims data. Demographics included as covariates in the models including age, sex, and commercial vs. Medicaid health insurance were determined at the index date. U.S. region and race were not included in statistical analyses, since geography was only available for commercially insured patients and race was only available for Medicaid patients.

### Statistical Analyses

An analysis of all pooled antipsychotic drugs as well as an analysis of prolactin category specific (1-3) exposures were performed. Bivariable analyses of the association between average exposure to antipsychotics and risk of invasive breast cancer were performed using Cox proportional hazards models. The primary exposure of antipsychotic medication utilization was modeled as described above by the average DDD, with the hazard ratio (HR) quantifying the risk associated with a 1.00-unit increase in the DDD.

Personal history of breast cancer was a censoring event during follow up through 30 days after an invasive breast cancer diagnosis, since coding of personal history in a short time frame after diagnosis likely indicates prior malignancy. Additional censoring, as described above, was performed at the earliest date of the following: death date, end of enrollment, or six years after the index date. Total follow-up was truncated at six years because of the small number of women remaining in the population. Three multivariable Cox proportional hazards models were used to adjust for known breast cancer risk factors that were determined *a priori* and included in the models regardless of significance. One model adjusted for known risk factors for breast cancer (e.g., obesity, hormone replacement therapy), another adjusted for age alone (age modeled as a cubic spline), and the third model adjusted for age and known risk factors for breast cancer. In addition to primary models, a sensitivity analysis was performed including additional adjustment for mental health conditions in the model with known risk factors for breast cancer, to account for potential confounding by severity of mental illness. Identical models were developed stratified by age less than or equal to or greater than 50 years as additional sensitivity analyses. We assessed ascertainment bias by counting the number of office/outpatient evaluation and management (E&M) encounters to providers other than psychiatrists/ psychologists during the baseline period for women with and without breast cancer. All data management and analyses were performed using SAS version 9.4 (SAS Institute Inc., Cary, NC) (38).

## Results

There were 1,248,951 commercially insured women from 2006-2016 with a median age of 42 years, and 313,888 Medicaid insured women from 2011-2016 with a median age of 37 years. Demographic characteristics of the 1,562,839 commercially and Medicaid-insured women who were users of antipsychotics, anticonvulsants and/ or lithium and met inclusion criteria for the study are in Table 1. In both cohorts, the median duration of total health insurance enrollment was 1,035 days. Overall, during the observation period of enrollment in the MarketScan Databases, 2,708 (0.2%) women were newly diagnosed with invasive breast cancer. Slightly over 40% of the women resided in the Southern U.S., consistent with database geographic representation. In the Medicaid database, approximately 60% of the women on study medications were White.

**Table 1.**
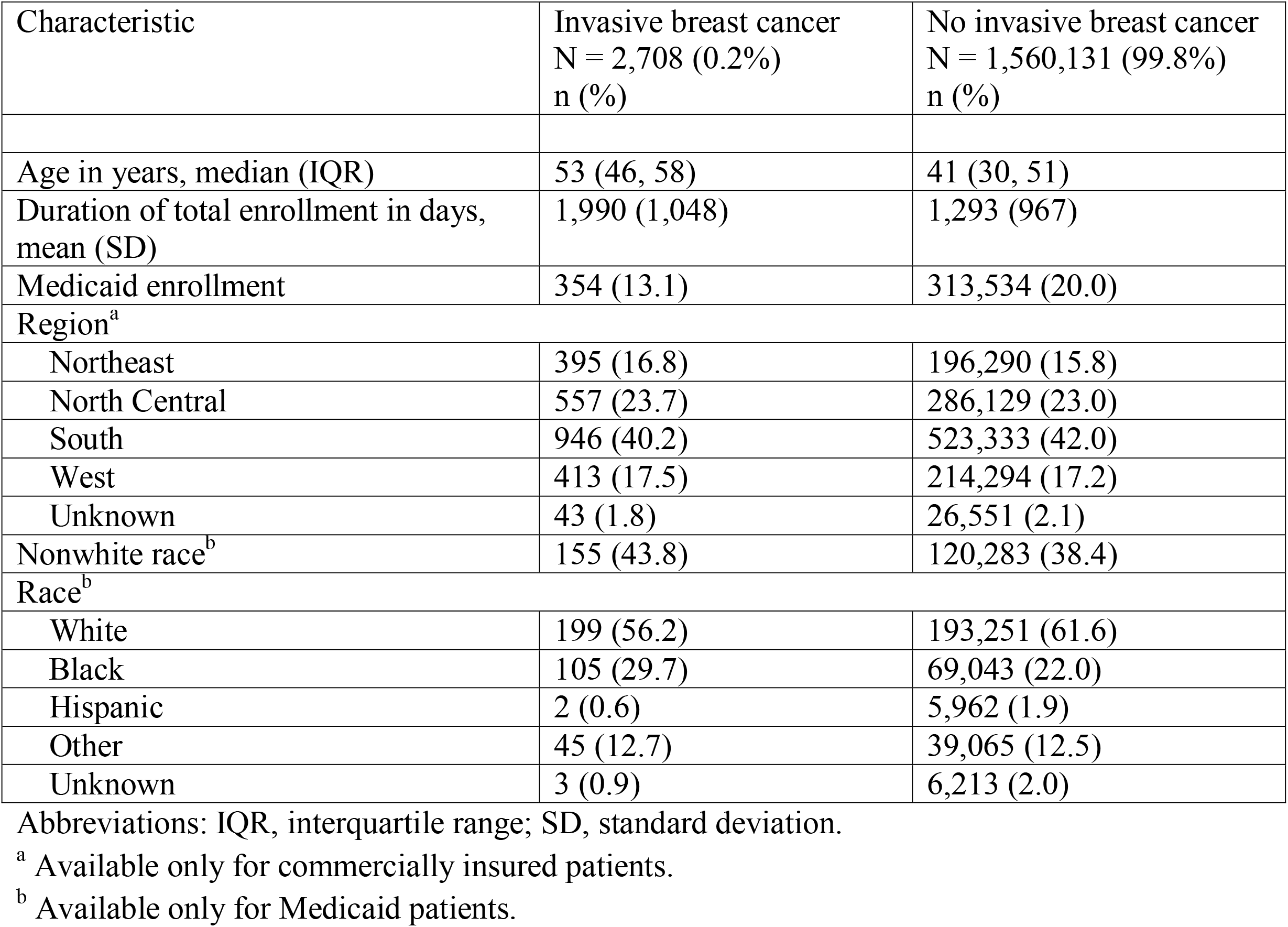
Demographics of the Population of Women Aged 18-64 years with Filled Prescriptions for Antipsychotic, Anticonvulsant, and/or Lithium Medications With and Without a New Diagnosis of Invasive Breast Cancer

| Characteristic | Invasive breast cancer<br>N = 2,708 (0.2%)<br>n (%) | No invasive breast cancer<br>N = 1,560,131 (99.8%)<br>n (%) |
| --- | --- | --- |
| Age in years, median (IQR) | 53 (46, 58) | 41 (30, 51) |
| Duration of total enrollment in days,<br>mean (SD) | 1,990 (1,048) | 1,293 (967) |
| Medicaid enrollment | 354 (13.1) | 313,534 (20.0) |
| Region<sup>a</sup> |  |  |
| Northeast | 395 (16.8) | 196,290 (15.8) |
| North Central | 557 (23.7) | 286,129 (23.0) |
| South | 946 (40.2) | 523,333 (42.0) |
| West | 413 (17.5) | 214,294 (17.2) |
| Unknown | 43 (1.8) | 26,551 (2.1) |
| Nonwhite race<sup>b</sup> | 155 (43.8) | 120,283 (38.4) |
| Race<sup>b</sup> |  |  |
| White | 199 (56.2) | 193,251 (61.6) |
| Black | 105 (29.7) | 69,043 (22.0) |
| Hispanic | 2 (0.6) | 5,962 (1.9) |
| Other | 45 (12.7) | 39,065 (12.5) |
| Unknown | 3 (0.9) | 6,213 (2.0) |

Abbreviations: IQR, interquartile range; SD, standard deviation.
<sup>a</sup> Available only for commercially insured patients.
<sup>b</sup> Available only for Medicaid patients.

Utilization of medications by category of antipsychotics, anticonvulsants and lithium is summarized in Table 2. Of the women with invasive breast cancer, 47% filled at least one prescription for a category 3 antipsychotic agent, while 21% filled at least one prescription for a category 1 agent. Of the women who were not diagnosed with invasive breast cancer during their period of study observation, approximately 50% filled at least one prescription for a category 3 antipsychotic agent, while 19% filled at least one prescription for a category 1 agent. In both groups over 50% of women filled at least one prescription for an anticonvulsant medication during their period of study observation. The median average DDD exposure to the three categories of antipsychotic medication was low, reflecting intermittent utilization of these medications (Table 2 and Appendix Figure, showing histograms of the average DDD for the 3 categories of antipsychotic medications for women with and without breast cancer).

**Table 2.** Utilization of Antipsychotic, Anticonvulsant, and Lithium Medications Among Adult Women Aged 18-64 years in the MarketScan Commercial and Multi-State Medicaid Databases

| Medication Category Based on propensity to elevate prolactin (category 1=highest, category 3=lowest) | Invasive breast cancer<br>N = 2,708<br>(0.2%) | No invasive breast cancer<br>N = 1,560,131 (99.8%) |
| --- | --- | --- |
| **Use of medication (1 or more fills), n (%)<sup>a</sup>** |  |  |
| Antipsychotic category 1 | 578 (21.3) | 290,976 (18.7) |
| Antipsychotic category 2 | 257 (9.5) | 164,369 (10.5) |
| Antipsychotic category 3 | 1,277 (47.2) | 777,211 (49.8) |
| Anticonvulsants | 1,394 (51.5) | 866,768 (55.6) |
| Lithium | 244 (9.0) | 136,840 (8.8) |
| **Average daily DDD, median (IQR)<sup>b</sup>** |  |  |
| Antipsychotic category 1 | 0.08 (0.01, 0.33) | 0.05 (0.01, 0.19) |
| Antipsychotic category 2 | 0.17 (0.04, 0.51) | 0.12 (0.03, 0.40) |
| Antipsychotic category 3 | 0.12 (0.03, 0.35) | 0.11 (0.03, 0.32) |
| Antipsychotic category 1–3 | 0.15 (0.04, 0.43) | 0.12 (0.03, 0.37) |

<sup>a</sup> Percentages total more than 100% because women could be counted in > 1 category.
<sup>b</sup> Average daily defined daily dose (DDD) = the cumulative DDD/total days of observed treatment period for a woman, per antipsychotic category.

Bivariable comparison of factors during the baseline period associated with invasive breast cancer among women in the cohort are summarized in Tables 2 and 3. Obesity, diabetes, hormone replacement therapy (in women 50 years and older, or younger women coded for surgical menopause), and preexisting benign breast disease were significantly more common in women newly diagnosed with invasive breast cancer than in women not diagnosed with breast cancer during the time period of their health insurance enrollment. The median age of women with breast cancer was 12 years older than women not diagnosed with breast cancer (53 vs 41 years, respectively, *p* < 0.0001). Although the number of outpatient E&M encounters in the baseline period was significantly different, the median number and interquartile ranges were the same in women with and without breast cancer (median 3 visits, IQR 1-6). In addition, over 80% of women in both groups had at least one E&M encounter during the baseline period.

**Table 3.** Bivariate Analysis of Factors During the Baseline Period Associated with Invasive Breast Cancer in Women with at least One Filled Prescription for Antipsychotic, Anticonvulsant, and/or Lithium Medications in the MarketScan Commercial and Multi-State Medicaid Databases

| Characteristic | Invasive Breast Cancer<br>N = 2,708 (0.1)<br>n (%) | No Breast Cancer<br>N = 1,560,131 (99.9)<br>n (%) | *P* |
| --- | --- | --- | --- |
| Age in years, median (IQR) | 53 (46, 58) | 41 (30, 51) | <.0001 |
| Number of Evaluation & Management<sup>a</sup> encounters during baseline period, median (IQR) | 3 (1, 6) | 3 (1, 6) | <.0001 |
| At least 1 Evaluation & Management encounter during baseline (%) | 2,290 (84.6) | 1,266,234 (81.2) | <.0001 |
| Mental health conditions |  |  |  |
| Bipolar disorder | 340 (12.6) | 245,069 (15.7) | <.0001 |
| Schizophrenia | 141 (5.2) | 60,677 (3.9) | 0.0004 |
| Major depression | 800 (29.5) | 472,411 (30.3) | 0.40 |
| Medicaid enrollment | 354 (13.1) | 313,534 (20.0) | <.0001 |
| Hormone replacement therapy<sup>b</sup> | 375 (13.8) | 82,003 (5.3) | <.0001 |
| Estrogen/progesterone-based HRT | 67 (2.5) | 10,549 (0.7) | <.0001 |
| Estrogen-only HRT | 308 (11.4) | 71,454 (4.6) | <.0001 |
| Obesity | 1,289 (47.6) | 573,689 (36.8) | <.0001 |
| Diabetes | 338 (12.5) | 137,312 (8.8) | <.0001 |
| Alcohol abuse | 77 (2.8) | 70,204 (4.5) | <.0001 |
| Preexisting benign breast disease | 59 (2.2) | 18,684 (1.2) | <.0001 |

Abbreviations: IQR, interquartile range; SD, standard deviation.
<sup>a</sup> Evaluation and management encounters include office/outpatient clinic or consultation visits.
<sup>b</sup> Hormone replacement therapy assessed only in women 50 years and older or in younger women with evidence of surgical menopause.

### Multivariable and Bivariable Models for Risk of Invasive Breast Cancer

Figure 1 provides the results of the bivariable and multivariable analyses of invasive breast cancer in women treated with pooled antipsychotic drugs (categories 1-3) compared to women treated only with anticonvulsants/ lithium (for full model results see Supplemental Table 3). Antipsychotic treated women had a slightly higher overall risk of breast cancer than control (only anticonvulsant/ lithium) users (unadjusted HR, 1.14; 95% CI, 1.07-1.21), with the hazards ratio corresponding to a one-unit increase in the average daily DDD for antipsychotic agents. There was little change in the risk of breast cancer associated with exposure to the combined categories of antipsychotics after adjustment for the known breast cancer risk factors enumerated in Table 3 (without age, adjusted HR 1.18; 95% CI, 1.11-1.25). The risk of breast cancer associated with exposure to the antipsychotic agents was non-significant after adjustment for age alone (adjusted HR 1.04; 95% CI, 0.97-1.12), and was nominally significant but slightly lower after adjustment for age plus known risk factors (adjusted HR 1.08; 95% CI, 1.01-1.16). The HRs for breast cancer associated with the pooled antipsychotic agents were similar in the sensitivity analyses stratified by age (Supplemental Tables 4 and 5).

**Figure 1.**
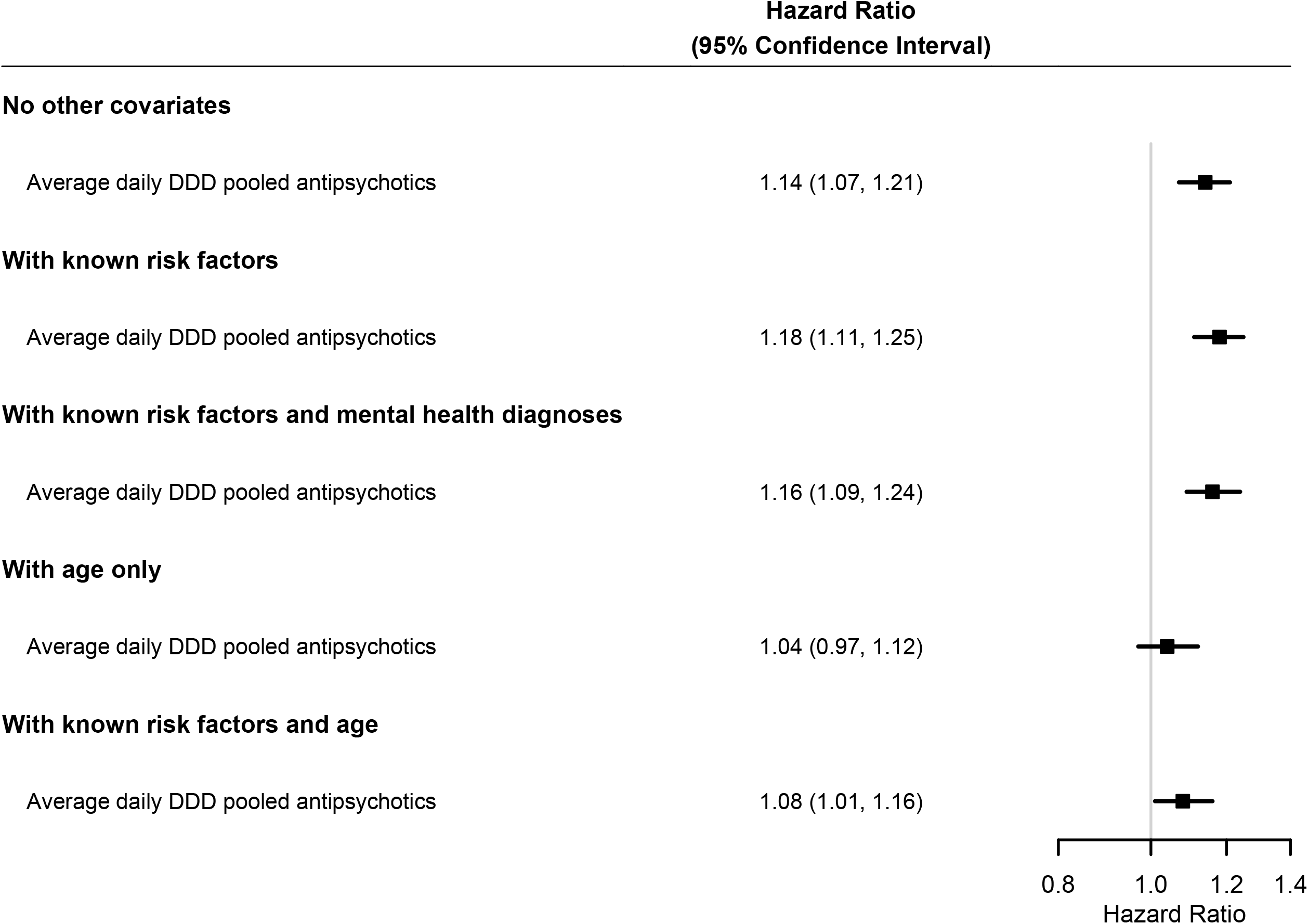
Bivariate and Multivariable Analyses of Risk of Invasive Breast Cancer in Women Treated with Pooled Antipsychotic Agents, Compared to Women Treated Only with Anticonvulsants or Lithium. Known risk factors for breast cancer included in the model were as follows: estrogen/progesterone-based hormone replacement therapy, estrogen-only hormone replacement therapy, diabetes, obesity, alcohol abuse, and preexisting benign breast disease. Medicaid enrollment was included in the model as a proxy for younger age at first birth and parity. Mental health diagnoses included were bipolar disorder, schizophrenia, and major depression. When noted, age was included using a cubic spline with seven knots. The HR represents the increase in breast cancer risk per one unit of the average defined daily dose (DDD) of the pooled antipsychotic medications.

Figure 2 shows results of analyses considering exposure to antipsychotic categories 1–3 individually (for full model results see Supplemental Table 6). In bivariable analysis, category one (high prolactin drugs) was associated with significantly increased risk (HR 1.24; 95% CI, 1.14-1.36), category two (mid prolactin) and category three (low prolactin) drugs were not associated with significantly increased risk of breast cancer. The results were similar when adjusted for known risk factors: category one (adjusted HR 1.30; 95% CI 1.20-1.40), category 2 (adjusted HR 1.18; 95% CI 1.01-1.37), and category 3 drugs (adjusted HR 1.07; 95% CI 0.96-1.19). The significant association of category 1 drugs (highest propensity to elevate prolactin) remained after adjustment for known risk factors and mental health conditions (adjusted HR 1.27; 95% CI 1.17-1.39) and after adjustment for age plus known risk factors (adjusted HR 1.23; 95% CI 1.11-1.35), while the category 2 and 3 drugs were not associated with increased risk of breast cancer. The results were similar in the sensitivity analyses restricted to women aged 50 and younger and women older than 50 years (Supplemental Tables 7 and 8).

**Figure 2.**
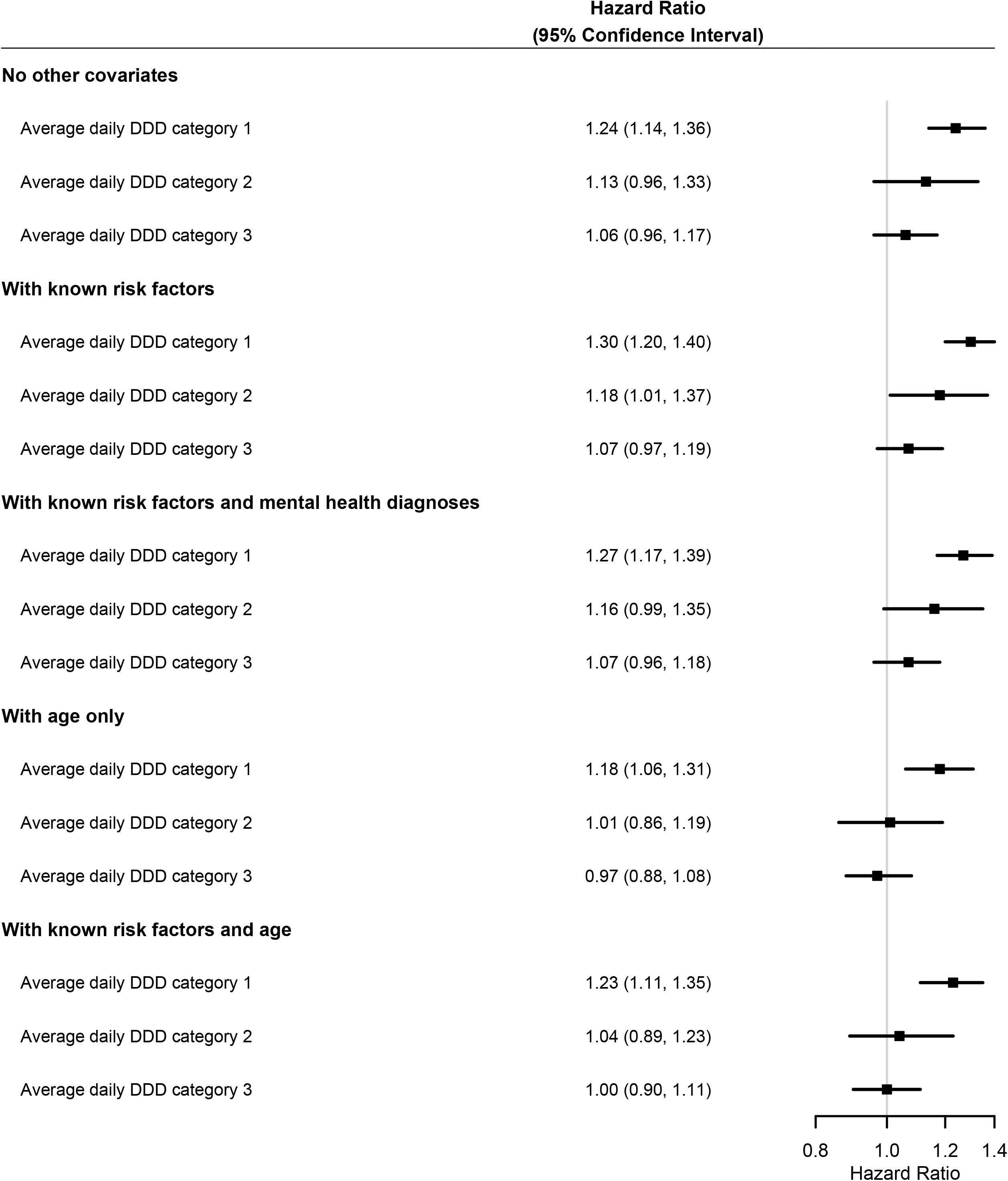
Bivariate and Multivariable Analyses of Risk of Invasive Breast Cancer in Women Treated with Antipsychotic Agents by Category of Propensity to Elevate Prolactin, Compared to Women Treated Only with Anticonvulsants or Lithium. Known risk factors for breast cancer included in the model were as follows: estrogen/progesterone-based hormone replacement therapy, estrogen-only hormone replacement therapy, diabetes, obesity, alcohol abuse, and preexisting benign breast disease. Medicaid enrollment was included in the model as a proxy for younger age at first birth and parity. Mental health diagnoses included were bipolar disorder, schizophrenia, and major depression. When noted, age was included using a cubic spline with seven knots. The HR represents the increase in breast cancer risk per one unit of the average defined daily dose (DDD) of the individual category of antipsychotic medications.

## Discussion

In the largest observational study evaluating risk of breast cancer in U.S. women taking antipsychotic drugs, we observed that women who filled prescriptions for prolactin-elevating category 1 antipsychotic drugs had a significantly elevated 23% increased risk of breast cancer per one-unit increase in the defined daily dose, compared to women with prescriptions only for comparator medications (anticonvulsants and/or lithium), after adjusting for age and other risk factors. That is, a patient on two-times the typical dose of a category one-antipsychotic would experience a 23% higher risk than a patient on the standard accepted dose. However, we also note that, in the population studied here, very few women were exposed to doses of antipsychotics that would result in meaningfully elevated risk.

We controlled for accepted risk factors for invasive breast cancer available in the administrative claims data, and used a rigorous approach to model exposure to the drugs by standardizing all drugs to DDD and calculating the average daily exposure to the individual drugs over the observed treatment period. We did not attempt to distinguish between new and prevalent users, because the medications are typically used both long-term or episodically. Therefore, we used the average daily drug category exposure during the study observation period as a proxy for lifetime exposure. Important imitations of our study include residual confounding due to parity, age at first birth, and race, which were not available. Also, the analysis population could not include women age 65 and older due to database limitations. Although the number of outpatient evaluation and management visits in the baseline period were the same in women with and without breast cancer, the lack of clinical detail does not allow us to rule out some degree of ascertainment bias. Mitigating these limitations, this studied utilized three separate antipsychotic categories and an active comparator control group (treated with only anticonvulsants and/or lithium). Only category one drugs were associated with significant risk, with the two other drug categories having no association with increased risk of breast cancer. When pooled together, the observed small risk of breast cancer associated with antipsychotics is likely attributable to the differential effects of category one drugs. The data are consistent with prior studies that suggest the possibility that antipsychotic drugs contribute to breast cancer risk, but also builds on prior research by suggesting that this risk may be limited to, or stronger for drugs with a high propensity to elevate prolactin (2-4, 6). The main strength if this study is analyzing risk by first separating drugs by prolactin elevating categories.

### Clinical Implications

Mentally ill patients face disparities in screening/ prevention of common cancers and are more likely to die from it. Women with psychotic disorders are half as likely as the general population to receive mammography screening (40). Our data and other publications (3,4,11,17) suggest that the use of category one antipsychotic drugs may contribute to incident cases of breast cancer. However, any risk must be weighed against potential benefits. Antipsychotic drugs have life-saving properties and should not be avoided due to potential risk for breast cancer (10). Our findings contribute to knowledge of the safety profile of antipsychotic medications and may be useful to patients considering use of these medications. Further prospective studies aimed at studying predictive biomarkers (prolactin levels, prolactin gene expression and mammographic breast density) are needed to design appropriate intervention studies for women taking antipsychotic drugs. In the meantime, we recommend vigilance to mitigate breast cancer risk in category one treated patients by monitoring serum prolactin levels, particularly in those treated with category one drugs (40). Moreover, women aged 40 and older taking category 1 drugs may be considered for mammography screening. Hyperprolactinemia can be avoided by lowering antipsychotic doses, switching antipsychotics, or by adding aripiprazole or dopamine agonists (6).

## Data Availability

None

## ACKNOWLEGMENTS

IBM Watson Health and MarketScan are trademarks of IBM Corporation in the United States, other countries or both. Support for this study was provided by an award from the Alvin J. Siteman Cancer Center. Programming and analysis for this study was conducted by the Center for Administrative Data Research, supported in part by the Washington University Institute of Clinical and Translational Sciences grant UL1 TR002345 from the National Center for Advancing Translational Sciences (NCATS) of the National Institutes of Health (NIH), Grant Number R24 HS19455 through the Agency for Healthcare Research and Quality (AHRQ).

