## Supplemental Tables for "Antipsychotic Drugs and the Risk of Breast Cancer"

**Supplemental Table 1. Antipsychotic, Anticonvulsant, and Lithium Medications Used to Define the Study Cohort Based on Category of Prolactin Elevation, and Maximum Defined Daily Dose Limits Allowed for Each Antipsychotic Medication**

| Category of prolactin elevation | Medication Class | Generic name | Maximum Defined Daily Dose Limit (DDD)^a^ |
| --- | --- | --- | --- |
| 1 | Antipsychotic | chlorpromazine | 10 |
|  |  | fluphenazine | 8 |
|  |  | haloperidol | 12.5 |
|  |  | loxapine | 3 |
|  |  | molindone | 4.5 |
|  |  | paliperidone | 5 |
|  |  | perphenazine | 2.133 |
|  |  | pimozide | 6 |
|  |  | prochlorperazine^b^ | 1.5 |
|  |  | risperidone | 1.6 |
|  |  | thioridazine | 3.333 |
|  |  | trifluoperazine | 3.25 |
| 2 | Antipsychotic | iloperidone | 2 |
|  |  | lurasidone | 2.667 |
|  |  | olanzapine | 6 |
| 3 | Antipsychotic | aripiprazole | 2 |
|  |  | asenapine | 1 |
|  |  | brexpiprazole | 4 |
|  |  | cariprazine | 4 |
|  |  | clozapine | 6.667 |
|  |  | quetiapine | 3 |
|  |  | ziprasidone | 4 |
| 4 | Anticonvulsants | carbamazepine | N/A |
|  |  | lamotrigine | N/A |
|  |  | oxcarbazepine | N/A |
|  |  | valproate | N/A |
| 5 | Lithium | lithium | N/A |

Abbreviation: N/A, not applicable

^a^ The maximum Daily Defined Dose (DDD) limit was established by the senior author.

^b^ Prochloperazine was not included as an inclusion criterion for antipsychotic exposure, but was used to calculate cumulative exposure to category 1 medications among women meeting criteria for study inclusion due to other category 1-5 medication exposure(s).

To calculate drug exposure for individuals in the dataset, for each individual prescription drug claim, the drug strength and metric quantity were used to compute the total amount of drug in milligrams (millimoles for lithium). This total amount was divided by the days’ supply to obtain the milligrams per day of the individual drug. The WHO generic name-specific conversion factors were then used to calculate the DDDs per day.^1^ The values were compared to the maximum DDD limits in the table above, and any values exceeding the limit were reset to the maximum allowable DDD.

Women were allowed to have more than one prescription at the same time, including of the same medication. This could be due to early refilling of medications, adjustment of dosage, and other factors.

1. World Health Organization. 2020. Collaborating centre for drug statistics methodology. Guidelines for ATC classification and DDD assignment. <https://www.whocc.no/atc_ddd_index/>

**Supplemental Table 2. Codes and Medications to Identify Invasive Breast Cancer, Benign Breast Disease, and Covariates of Interest**

| Category | ICD-9-CM/ICD-10 Diagnosis Codes | ICD-9-CM/ICD-10-PCS Procedure Codes | CPT-4/Revenue Center Codes | Medications (generic name or HCPCS code) |
| --- | --- | --- | --- | --- |
| Invasive Breast Cancer | 174.0–174.9, 198.2  C50.011-C50.119, C50.111-C50.119, C50.211-C50.219, C50.311-C50.319, C50.411-C50.419, C50.511-C50.519, C50.811-C50.819, C50.911-C50.919 |  |  |  |
| Breast Carcinoma *in situ* | 233.0  D05.10-D05.92 |  |  |  |
| Pathology microscopic examination of tissue |  |  | 88302-88388 |  |
| Mastectomy |  | 85.33-85.36, 85.41-85.48  0HTT0ZZ, 0HTU0ZZ, 0HTV0ZZ | 19303-19307 |  |
| Breast-conserving surgery |  | 85.20-85.23  0HBT0ZX, 0HBT0ZZ, 0HBT7ZX, 0HBT7ZZ, 0HBT8ZX, 0HBT8ZZ, 0HBU0ZX, 0HBU0ZZ, 0HBU7ZX, 0HBU7ZZ, 0HBU8ZX, 0HBU8ZZ, 0HBV0ZX, 0HBV0ZZ, 0HBV7ZX, 0HBV7ZZ, 0HBV8ZX, 0HBV8ZZ, 0HTWXZZ, 0HTXXZZ | 19120, 19125, 19126, 19160, 19162, 19301, 19302 |  |
| Benign Breast Disease | 610.0-610.3, 610.8, 610.9, 217, 238.3, 239.3, 611.72  N60.01-N60.39, N60.81-N60.99, N63,  D24.1-D24.9, D48.60-D48.62, D49.3 |  |  |  |
| Breast biopsy |  | 85.11, 85.12  0HBT3ZX, 0HBT3ZZ, 0HBU3ZX, 0HBU3ZZ, 0HBV3ZX, 0HBV3ZZ, 0HBW3ZX, 0HBW3ZZ, 0HBX3ZX, 0HBX3ZZ, 0HBW0ZX, 0HBW0ZZ, 0HBW7ZX, 0HBW7ZZ, 0HBW8ZX, 0HBW8ZZ, 0HBWXZX, 0HBWXZZ, 0HBX0ZX, 0HBX0ZZ, 0HBX7ZX, 0HBX7ZZ, 0HBX8ZX, 0HBX8ZZ, 0HBXXZX, 0HBXXZZ | 19081, 19082, 19083, 19084, 19085, 19086, 19100, 19101, 19102, 19103 |  |
| Chemotherapy |  | 99.25  3E03005, 3E03305, 3E04005, 3E04305, 3E05005, 3E05305, 3E06005, 3E06305 | Revenue center: 0331, 0332, 0335 | 96400, 96401, 96403-96440  J9000-J9999, Q0083-Q0085 |
| Obesity | 278.00, 278.01 278.03, 649.10- 649,14, 793.91, V85.30-V85.45, V85.54  E66.01, E66.09, E66.1, E66.2, E66.8, E66.9, O99.210- O99.215, R93.9, Z68.30- Z68.45, Z68.54 |  |  |  |
| Obesity Proxies (sleep apnea, gastroesophageal reflux disease, lipodystrophy, other lipidemias) | 327.23, G47.33,  530.81, K21.9,  272.6, E88.1,  272.0- 272.5, 272.7-272.9,  E75.21, E75.22, E75.29, E77.0, E77.1, E78.00-E78.6, E78.81-E78.9, E88.89 |  |  | Therapeutic class Antihyperlipidemic medications (statins),  amlodipine/atorvastatin,  colestipol, cholestyramine, colesevelam, ezetimibe, fenofibrate, gemfibrozil, niacin, omega-3-acid ethyl esters, icosapentethyl |
| Diabetes | 249.00-249.91, 250.00-250.93, 648.00-648.04, 775.1  E08.00-E13.9 O24.011-O24.33, O24.811-O24.93 |  |  | acarbose,  acetohexamide albiglutide, alogliptin, bromocriptine, canagliflozin, chlorpropamide, dapagliflozin, dulaglutide, empagliflozin, exenatide, gliclazide glimepiride, glipizide, glyburide,  insulin, linagliptin, liraglutide, metformin, miglitol, nateglinide, pioglitazone, pramlintide, repaglinide, rosiglitazone, saxagliptin, sitagliptin, tolazamide, tolbutamide, troglitazone |
| Smoking/smoking cessation therapy | 305.1, 649.00-649.04, V15.82,  F17.200, O99.330- O99.335 |  |  | varenicline |
| Smoking-related diseases | 162.2-162.9 231.1, C34.00-C34.92, C7A.090, D02.20-D02.22, Z85.118,  410.00- 412,  I21.01- I22.9, I23.8, I25.2,  413.0-413.9, I20.0-I20.9,  411.0-411.89, 414.8, 414.9, I24.0-I24.9, I25.5 I25.89, I25.9,  431, 432.9, 433.00-438.89,  I61.0-I61.9, I62.9, I63.00-I63.9, I65.01- I66.9, I67.1, I67.2, I67.4, I67.5, I67.7 I67.81, I67.82, I67.89, I69.10-I69.198, I69.30-I69.398, I69.90-I69.998, G45.0- G45.2, G45.4, G45.8, G45.9,  491.20, 491.21 496, J44.0, J44.1, J44.9 |  |  |  |
| Alcohol abuse | 291.0-291.3, 291.5, 291.8, 291.81, 291.82, 291.89, 291.9, 303.00-303.93, 305.00-305.03,  F10.10-F10.29, F10.921, F10.94, F10.950 -F10.99 |  |  |  |
| Hormone replacement therapy* |  |  |  | Therapeutic class - estrogens & combinations |
| Acquired absence of genital organs | V45.77, Z90.710-Z90.712, Z90.722 |  |  |  |
| Evaluation and management (E&M) encounters |  |  | 99201-99205, 99211-99215, 99241-99245, 99385, 99386, 99395, 99396 |  |
| *restricted to women aged >= 50 years or younger women with a diagnosis of acquired absence of genital organs. Combinations with progesterone were considered separately from estrogen-only containing prescriptions. | | | | |

**Supplemental Table 3. Bivariable and Multivariable Analyses of Risk of Invasive Breast Cancer in Women Treated With Pooled Antipsychotic Agents, Compared to Women Treated Only With Anticonvulsants and/or Lithium**

|  | No other covariates | With known risk factors | With known risk factors plus mental health diagnoses | With age alone^a^ | With known risk factors + age^a^ |
| --- | --- | --- | --- | --- | --- |
| Variable | HR (95% CI) | HR (95% CI) | HR (95% CI) | HR (95% CI) | HR (95% CI) |
| Average daily DDD category 1-3 drugs^b^ | 1.14 (1.07-1.21) | 1.18 (1.11-1.25) | 1.16 (1.09-1.24) | 1.04 (0.97-1.12) | 1.08 (1.01-1.16) |
| Medicaid enrollment |  | 0.57 (0.51-0.64) | 0.55 (0.49-0.62) |  | 0.77 (0.68-0.86) |
| Estrogen/progesterone-based hormone replacement therapy |  | 2.33 (1.82-2.97) | 2.32 (1.82-2.96) |  | 1.43 (1.12-1.83) |
| Estrogen-only hormone replacement therapy |  | 1.85 (1.64-2.09) | 1.85 (1.64-2.09) |  | 1.03 (0.91-1.16) |
| Diabetes |  | 1.24 (1.11-1.39) | 1.23 (1.10-1.38) |  | 1.02 (0.91-1.15) |
| Obesity |  | 1.48 (1.37-1.61) | 1.48 (1.37-1.61) |  | 0.98 (0.91-1.07) |
| Alcohol abuse |  | 0.82 (0.66-1.03) | 0.83 (0.66-1.05) |  | 0.86 (0.69-1.08) |
| Preexisting benign breast disease |  | 1.47 (1.14-1.91) | 1.47 (1.14-1.91) |  | 1.33 (1.03-1.72) |
| Bipolar disorder |  |  | 0.87 (0.78-0.98) |  |  |
| Schizophrenia |  |  | 1.28 (1.06-1.54) |  |  |
| Major depression |  |  | 0.98 (0.90-1.06) |  |  |

^a^ Age included in the model using a cubic spline with 7 knots.

^b^ Increase in risk per average daily DDD. Specifically, the HR is the increase in breast cancer risk per one defined daily dose in average daily exposure to all category 1-3 antipsychotics.

**Supplemental Table 4. Bivariable and Multivariable Analyses of Risk of Invasive Breast Cancer in Women Between 18 and 50 Years of Age Treated With Pooled Antipsychotic Agents, Compared to Women Treated Only With Anticonvulsants and/or Lithium**

|  | No other covariates | With known risk factors | With known risk factors plus mental health diagnoses | With age alone^a^ | With known risk factors + age^a^ |
| --- | --- | --- | --- | --- | --- |
| Variable | HR (95% CI) | HR (95% CI) | HR (95% CI) | HR (95% CI) | HR (95% CI) |
| Average daily DDD category 1-3 drugs^b^ | 1.12 (1.01-1.25) | 1.16 (1.05-1.29) | 1.14 (1.02-1.27) | 1.01 (0.90-1.14) | 1.05 (0.93-1.18) |
| Medicaid enrollment |  | 0.55 (0.46-0.66) | 0.54 (0.45-0.64) |  | 0.75 (0.63-0.89) |
| Estrogen/progesterone-based hormone replacement therapy |  | 7.13 (3.52-14.45) | 7.13 (3.52-14.44) |  | 3.59 (1.77-7.23) |
| Estrogen-only hormone replacement therapy |  | 1.04 (0.57-1.90) | 1.04 (0.57-1.90) |  | 0.54 (0.29-0.98) |
| Diabetes |  | 1.13 (0.91-1.40) | 1.13 (0.91-1.39) |  | 0.98 (0.79-1.21) |
| Obesity |  | 1.35 (1.18-1.54) | 1.35 (1.19-1.54) |  | 0.98 (0.86-1.12) |
| Alcohol abuse |  | 0.78 (0.55-1.12) | 0.79 (0.56-1.13) |  | 0.75 (0.53-1.07) |
| Preexisting benign breast disease |  | 1.46 (0.93-2.31) | 1.47 (0.93-2.31) |  | 1.10 (0.70-1.74) |
| Bipolar disorder |  |  | 0.91 (0.77-1.08) |  |  |
| Schizophrenia |  |  | 1.33 (0.98-1.80) |  |  |
| Major depression |  |  | 0.97 (0.85-1.10) |  |  |

^a^ Age included in the model using a cubic spline with 5 knots.

^b^ Increase in risk per average daily DDD. Specifically, the HR is the increase in breast cancer risk per one defined daily dose in average daily exposure to all category 1-3 antipsychotics.

**Supplemental Table 5. Bivariable and Multivariable Analyses of Risk of Invasive Breast Cancer in Women Between 51 and 64 Years of Age Treated With Pooled Antipsychotic Agents, Compared to Women Treated Only With Anticonvulsants and/or Lithium**

|  | No other covariates | With known risk factors | With known risk factors plus mental health diagnoses | With age alone^a^ | With known risk factors + age^a^ |
| --- | --- | --- | --- | --- | --- |
| Variable | HR (95% CI) | HR (95% CI) | HR (95% CI) | HR (95% CI) | HR (95% CI) |
| Average daily DDD category 1-3 drugs^b^ | 1.06 (0.97-1.15) | 1.10 (1.01-1.20) | 1.10 (1.00-1.21) | 1.06 (0.97-1.16) | 1.10 (1.01-1.20) |
| Medicaid enrollment |  | 0.76 (0.65-0.89) | 0.76 (0.65-0.90) |  | 0.79 (0.67-0.92) |
| Estrogen/progesterone-based hormone replacement therapy |  | 1.31 (1.01-1.70) | 1.31 (1.01-1.70) |  | 1.33 (1.03-1.73) |
| Estrogen-only hormone replacement therapy |  | 1.06 (0.94-1.21) | 1.06 (0.93-1.21) |  | 1.07 (0.94-1.22) |
| Diabetes |  | 1.07 (0.93-1.22) | 1.07 (0.93-1.22) |  | 1.04 (0.91-1.19) |
| Obesity |  | 1.04 (0.94-1.15) | 1.04 (0.94-1.15) |  | 0.99 (0.89-1.09) |
| Alcohol abuse |  | 0.92 (0.68-1.24) | 0.92 (0.68-1.24) |  | 0.96 (0.71-1.30) |
| Preexisting benign breast disease |  | 1.44 (1.05-1.98) | 1.44 (1.05-1.97) |  | 1.47 (1.07-2.01) |
| Bipolar disorder |  |  | 0.95 (0.82-1.11) |  |  |
| Schizophrenia |  |  | 1.00 (0.79-1.27) |  |  |
| Major depression |  |  | 1.02 (0.92-1.14) |  |  |

^a^ Age included in the model using a cubic spline with 5 knots.

^b^ Increase in risk per average daily DDD. Specifically, the HR is the increase in breast cancer risk per one defined daily dose in average daily exposure to all category 1-3 antipsychotics.

**Supplemental Table 6. Bivariable and Multivariable Analyses of Risk of Invasive Breast Cancer in Women Treated With Antipsychotic Agents by Category of Propensity to Elevate Prolactin, Compared to Women Treated Only With Anticonvulsants and/or Lithium**

|  | No other covariates | With known risk factors | With known risk factors plus mental health diagnoses | With age alone^a^ | With known risk factors and age^a^ |
| --- | --- | --- | --- | --- | --- |
| Variable | HR (95% CI) | HR (95% CI) |  |  | HR (95% CI) |
| Average daily DDD category 1 drugs (highest propensity to elevate prolactin)^b^ | 1.24 (1.14-1.36) | 1.30 (1.20-1.40) | 1.27 (1.17-1.39) | 1.18 (1.06-1.31) | 1.23 (1.11-1.35) |
| Average daily DDD category 2 drugs^b^ | 1.13 (0.96-1.33) | 1.18 (1.01-1.37) | 1.16 (0.99-1.35) | 1.01 (0.86-1.19) | 1.04 (0.89-1.23) |
| Average daily DDD category 3 drugs (lowest propensity to elevate prolactin)^b^ | 1.06 (0.96-1.17) | 1.07 (0.97-1.19) | 1.07 (0.96-1.18) | 0.97 (0.88-1.08) | 1.00 (0.90-1.11) |
| Medicaid enrollment |  | 0.56 (0.50-0.63) | 0.55 (0.49-0.62) |  | 0.76 (0.68-0.85) |
| Estrogen/progesterone-based hormone replacement therapy |  | 2.32 (1.82-2.97) | 2.32 (1.82-2.96) |  | 1.43 (1.12-1.83) |
| Estrogen-only hormone replacement therapy |  | 1.85 (1.64-2.09) | 1.85 (1.64-2.09) |  | 1.03 (0.91-1.16) |
| Diabetes |  | 1.24 (1.11-1.39) | 1.23 (1.10-1.39) |  | 1.02 (0.91-1.15) |
| Obesity |  | 1.49 (1.37-1.61) | 1.49 (1.37-1.61) |  | 0.99 (0.91-1.07) |
| Alcohol abuse |  | 0.82 (0.66-1.03) | 0.83 (0.66-1.05) |  | 0.86 (0.69-1.08) |
| Preexisting benign breast disease |  | 1.47 (1.13-1.90) | 1.47 (1.13-1.90) |  | 1.33 (1.02-1.72) |
| Bipolar disorder |  |  | 0.88 (0.78-0.99) |  |  |
| Schizophrenia |  |  | 1.26 (1.05-1.52) |  |  |
| Major depression |  |  | 0.98 (0.90-1.07) |  |  |

^a^ Age included in the model using a cubic spline with 7 knots.

^b^Increase in risk per average daily DDD. Specifically, the HR is the increase in breast cancer risk per one defined daily dose in average daily exposure to the individual category of antipsychotic medications.

**Supplemental Table 7. Bivariable and Multivariable Analyses of Risk of Invasive Breast Cancer in Women Between 18 and 50 Years of Age Treated With Antipsychotic Agents by Category of Propensity to Elevate Prolactin, Compared to Women Treated Only With Anticonvulsants and/or Lithium**

|  | No other covariates | With known risk factors | With known risk factors plus mental health diagnoses | With age alone^a^ | With known risk factors and age^a^ |
| --- | --- | --- | --- | --- | --- |
| Variable | HR (95% CI) | HR (95% CI) | HR (95% CI) | HR (95% CI) | HR (95% CI) |
| Average daily DDD category 1 drugs (highest propensity to elevate prolactin)^b^ | 1.19 (0.99-1.43) | 1.26 (1.07-1.47) | 1.22 (1.02-1.45) | 1.14 (0.93-1.40) | 1.19 (1.00-1.43) |
| Average daily DDD category 2 drugs^b^ | 1.16 (0.88-1.52) | 1.23 (0.94-1.59) | 1.20 (0.92-1.56) | 1.07 (0.81-1.42) | 1.12 (0.85-1.47) |
| Average daily DDD category 3 drugs (lowest propensity to elevate prolactin)^b^ | 1.06 (0.90-1.25) | 1.08 (0.91-1.27) | 1.07 (0.90-1.26) | 0.92 (0.78-1.10) | 0.95 (0.80-1.13) |
| Medicaid enrollment |  | 0.55 (0.46-0.65) | 0.53 (0.48-0.64) |  | 0.74 (0.62-0.88) |
| Estrogen/progesterone-based hormone replacement therapy |  | 7.15 (3.53-14.49) | 7.14 (3.53-14.47) |  | 3.60 (1.78-7.30) |
| Estrogen-only hormone replacement therapy |  | 1.04 (0.57-1.90) | 1.04 (0.57-1.90) |  | 0.54 (0.29-0.99) |
| Diabetes |  | 1.13 (0.92-1.40) | 1.13 (0.91-1.40) |  | 0.98 (0.79-1.21) |
| Obesity |  | 1.35 (1.19-1.54) | 1.35 (1.19-1.54) |  | 0.98 (0.86-1.12) |
| Alcohol abuse |  | 0.79 (0.55-1.12) | 0.79 (0.56-1.13) |  | 0.75 (0.53-1.07) |
| Preexisting benign breast disease |  | 1.46 (0.93-2.30) | 1.46 (0.93-2.31) |  | 1.10 (0.70-1.74) |
| Bipolar disorder |  |  | 0.91 (0.77-1.09) |  |  |
| Schizophrenia |  |  | 1.31 (0.97-1.78) |  |  |
| Major depression |  |  | 0.97 (0.85-1.11) |  |  |

^a^ Age included in the model using a cubic spline with 5 knots.

^b^Increase in risk per average daily DDD. Specifically, the HR is the increase in breast cancer risk per one defined daily dose in average daily exposure to the individual category of antipsychotic medications.

**Supplemental Table 8. Bivariable and Multivariable Analyses of Risk of Invasive Breast Cancer in Women Between 51 and 64 Years of Age Treated With Antipsychotic Agents by Category of Propensity to Elevate Prolactin, Compared to Women Treated Only With Anticonvulsants and/or Lithium**

|  | No other covariates | With known risk factors | With known risk factors plus mental health diagnoses | With age alone^a^ | With known risk factors and age^a^ |
| --- | --- | --- | --- | --- | --- |
| Variable | HR (95% CI) | HR (95% CI) | HR (95% CI) | HR (95% CI) | HR (95% CI) |
| Average daily DDD category 1 drugs (highest propensity to elevate prolactin)^b^ | 1.20 (1.06-1.36) | 1.25 (1.12-1.40) | 1.25 (1.12-1.40) | 1.20 (1.05-1.36) | 1.24 (1.11-1.40) |
| Average daily DDD category 2 drugs^b^ | 1.00 (0.81-1.22) | 1.03 (0.84-1.26) | 1.03 (0.85-1.27) | 0.98 (0.80-1.20) | 1.01 (0.83-1.24) |
| Average daily DDD category 3 drugs (lowest propensity to elevate prolactin)^b^ | 0.99 (0.87-1.13) | 1.02 (0.89-1.16) | 1.02 (0.89-1.17) | 1.00 (0.88-1.14) | 1.03 (0.90-1.18) |
| Medicaid enrollment |  | 0.75 (0.64-0.88) | 0.76 (0.64-0.89) |  | 0.78 (0.67-0.91) |
| Estrogen/progesterone-based hormone replacement therapy |  | 1.31 (1.01-1.70) | 1.31 (1.01-1.70) |  | 1.33 (1.03-1.73) |
| Estrogen-only hormone replacement therapy |  | 1.06 (0.94-1.21) | 1.06 (0.94-1.21) |  | 1.07 (0.94-1.22) |
| Diabetes |  | 1.07 (0.93-1.22) | 1.07 (0.93-1.22) |  | 1.04 (0.91-1.19) |
| Obesity |  | 1.04 (0.94-1.15) | 1.04 (0.94-1.15) |  | 0.99 (0.89-1.10) |
| Alcohol abuse |  | 0.92 (0.68-1.24) | 0.92 (0.68-1.24) |  | 0.96 (0.72-1.30) |
| Preexisting benign breast disease |  | 1.44 (1.05-1.98) | 1.44 (1.05-1.97) |  | 1.47 (1.07-2.01) |
| Bipolar disorder |  |  | 0.96 (0.83-1.13) |  |  |
| Schizophrenia |  |  | 0.99 (0.78-1.25) |  |  |
| Major depression |  |  | 1.03 (0.92-1.15) |  |  |

^a^ Age included in the model using a cubic spline with 5 knots.

^b^Increase in risk per average daily DDD. Specifically, the HR is the increase in breast cancer risk per one defined daily dose in average daily exposure to the individual category of antipsychotic medications.
